# Neonatal white matter tract microstructure and 2-year language outcomes after preterm birth

**DOI:** 10.1101/19011361

**Authors:** Sarah E. Dubner, Jessica Rose, Heidi M. Feldman, Katherine E. Travis

## Abstract

**Aim:** To determine whether variability in diffusion MRI (dMRI) white matter tract metrics, obtained in a cohort of preterm infants prior to neonatal hospital discharge, would be associated with language outcomes at age 2 years, after consideration of age at scan and number of major neonatal complications. Method: 30 children, gestational age 28.9 (2.4) weeks, underwent dMRI at mean post menstrual age 36.4 (1.4) weeks and language assessment with the Bayley Scales of Infant Development–III at mean age 22.2 (1.7) months chronological age. Mean fractional anisotropy (FA) and mean diffusivity (MD) were calculated for 5 white matter tracts. Hierarchical linear regression assessed associations between tract FA, moderating variables, and language outcomes. Results: FA of the left inferior longitudinal fasciculus accounted for 17% (p = 0.03) of the variance in composite language and FA of the posterior corpus callosum accounted for 19% (p = 0.02) of the variance in composite language, beyond that accounted for by post menstrual age at scan and neonatal medical complications. The number of neonatal medical complications moderated the relationship between language and posterior corpus callosum FA but did not moderate the association in the other tract. Conclusion: Language at 2 is associated with white matter metrics in early infancy in preterm children. The different pattern of associations by fiber group may relate to the stage of brain maturation and/or the nature and timing of medical complications related to preterm birth. Future studies should replicate these findings with a larger sample size to assure reliability of the findings.

## 1. INTRODUCTION*^1^*

Children born preterm (PT) are at risk for expressive and receptive language deficits (Barre et al., 2011; Foster-Cohen et al., 2007; van Noort-van der Spek et al., 2012). Language deficits in PT children increase risk for later reading problems, (Wocadlo and Rieger, 2007) which, in turn, are associated with adolescent health behaviors and long-term academic and occupational success (Connor et al., 2014; Sanders et al., 2009). Efforts to understand the neurobiological basis for early language development in PT children may suggest avenues of intervention to improve language skills.

PT infants are at risk for white matter alterations (Volpe, 2009). Between 24 and 32 weeks gestational age (GA), pre-oligodendrocytes, the immature progenitors of myelinating glial cells (ie., oligodendrocytes), are susceptible to oxidative stress from hypoxia, ischemia, and inflammation, often failing to mature or myelinate properly (Volpe, 2009). Periventricular regions are particularly sensitive to oxidative stress-related injuries (Volpe, 2009). Diffusion MRI (dMRI) has become a preferred method for assessing and characterizing the properties of white matter pathways in the brain (Basser and Pierpaoli, 1996; Feldman et al., 2010). Diffusion tensor imaging (DTI) is an analytic approach to dMRI data that generates metrics used to index white matter microstructure, including fractional anisotropy (FA) and mean diffusivity (MD). Within white matter voxels, increased FA and decreased MD are generally associated with favorable neurobiological factors, such as increased myelination and axonal density. However, dMRI metrics are also influenced by additional structural and tissue properties that reduce FA and/or increase MD, including the proportion of crossing fibers within a voxel, axonal size, and membrane permeability (Jones and Cercignani, 2010). Evidence for lower FA and/or higher MD in dMRI studies of PT neonates performed at term equivalent age (TEA) compared with children born full term (FT) (Ball et al., 2013; Thompson et al., 2011) is consistent with evidence for reduced myelination and axonal growth in animal models of PT birth (Volpe, 2009).

Studies have examined concurrent relations between language and white matter pathway metrics in school-aged PT and FT children (Dodson et al., 2018; Travis et al., 2016), but few studies have examined later language skills in relation to white matter metrics in the neonatal period. Many children obtain a battery of MRI scans, including dMRI, before they are discharged from the neonatal intensive care unit (NICU), which usually occurs near 35 – 45 weeks post-menstrual age, or PMA. Emerging evidence suggests that variability in white matter microstructure assessed from these pre-discharge dMRIs relates to later language development in PT children. FA in the left and right arcuate fasciculi measured at 39-46 weeks in children born preterm was found to be independently associated with language outcomes at 22 months (Salvan et al., 2017). In a longitudinal cohort of PT children imaged at TEA, 2- and 4-years of age, (Young et al., 2017) slower rate of change of MD and radial diffusivity (RD) in the left internal and external capsules, and slower rate of change of axial diffusivity (AD) in the left posterior thalamic radiation was associated with lower 4-year old language scores. Additional studies are required to confirm and expand on these associations.

PT birth-related illnesses, such as bronchopulmonary dysplasia, have been associated with white matter metrics (Dubner et al., 2019; Shim et al., 2012) and with language outcomes (Singer et al., 2001). The degree to which PT children experience neonatal medical complications varies substantially. Studies have yet to determine whether white matter microstructure may explain variance in language outcomes beyond medical complications of PT birth, especially those likely to induce hypoxic, ischemic or neuroinflammatory responses. Further, studies have not yet explored whether these complications may also moderate associations between pre-discharge measures of white matter microstructure and language outcomes. Examining these relations in detail is pertinent to determining whether white matter microstructure may be an important early biomarker for language outcomes.

The present study was designed to determine (1) if language outcomes at 2 years related to variability in white matter microstructure in language-related white matter tracts on pre-discharge dMRI scans in PT infants, beyond the influence of developmental age at scan and neonatal medical complications and (2) whether the number of medical complications moderated any effects. We obtained measures for FA and MD from specific white matter pathways commonly implicated in processing linguistic information (Hickok and Poeppel, 2007) and in tracts in which white matter metrics and language related measures have been associated in PT children, including a dorsal pathway, the Arc-L (Dodson et al., 2018; Travis et al., 2016) and two bilateral ventral pathways, the ILF-L and ILF-R, and the UF-L and UF-R (Beaulieu et al., 2005; Ben-Shachar et al., 2007; Bruckert et al., 2019a; Dodson et al., 2018; Travis et al., 2016, 2017; Vandermosten et al., 2012). Additionally, we examined fiber pathways traversing the occipital segment of the corpus callosum (CC-Occ) for three reasons: (1) Intrahemispheric connections are postulated to play an important role in language processing in infancy (Perani et al., 2011). (2) Studies have found relations between language-related skills and the posterior corpus callosum in both PT (Dubner et al., 2019) and FT children (Dougherty et al., 2007; Frye et al., 2008; Hasan et al., 2012; Huber et al., 2019; Odegard et al., 2009). (3) Finally, this region is susceptible to injuries from medical complications of PT birth (Back, 2015; Volpe, 2009). We assessed FA and MD given our previous work, which found these metrics to be related to neurocognitive outcomes in sample of older children born PT with and without history of neonatal conditions associated with adverse developmental outcomes, such as bronchopulmonary dysplasia, necrotizing enterocolitis, or sepsis (Dubner et al., 2019). We know that many postnatal factors, including socioeconomic status and number of words adults use with children influence language development (Adams et al., 2018; Feldman, 2019). Nonetheless, we reasoned that the status of white matter pathways after the neonatal hospitalization might be associated with later language development. Therefore, in this study, we hypothesized that (1) language would be associated with white matter metrics in all of the selected tracts, independent of PMA and the number neonatal medical complications; and (2) that the number of neonatal medical complications would moderate the association between white matter metrics and language because of the putative susceptibility of white matter in PT born infants to oxidative-stress related injuries.

## 2. MATERIALS AND METHODS

### 2.1 Participants

Children ≤ 1500 grams or ≤ 32 weeks gestation at birth from the Lucile Packard Children’s Hospital (LPCH) Stanford Neonatal Intensive Care Unit (NICU) born between January 2010 and December 2011 were eligible for this prospective study (Rose et al., 2015). Exclusion criteria included evidence of genetic disorders or congenital brain anomalies. 66 preterm infants underwent dMRI imaging near hospital discharge. Per LPCH NICU protocol, MRIs were obtained when infants were stable in an open crib, requiring no more than supplemental oxygen for respiratory support, and > 34 weeks PMA. Of these, n = 36 participants were excluded for several reasons: T1-weighted scan issues (n = 9), incomplete diffusion scans (n = 1), excessive head movement (n = 21), presence of parenchymal brain disease, including periventricular leukomalacia, on clinical MRI (n = 2), or missing language assessment (n = 3). The final sample was 30 participants. There were no significant differences between the 30 included and 36 excluded participants from the initial sample in GA (28.9 (2.4) vs 28.9 (2.4) weeks, t(64) = 0.05, *p* = 0.96), birthweight (1085 (281) vs 1095 (258) grams, t(64) = 0.14, *p* = 0.89), or PMA (36.4 (1.4) vs 36.6 (1.2) weeks, t(64) = 0.72, *p* = 0.49). In addition there were no differences between included and excluded participants in proportion of males (36.7% vs 38.9%, *p* = 0.85), or proportion with sepsis (3.3% vs 13.9%, *p* = 0.21), bronchopulmonary dysplasia (16.7% vs 33.3%, *p* = 0.16), necrotizing enterocolitis (3.3% vs 16.8%, *p* = 0.12), patent ductus arteriosus (48.3% vs 41.7%, *p* = 0.62), retinopathy of prematurity (36.7% vs 41.7%, *p* = 0.68), or intraventricular hemorrhage (20.0% vs 22.2%, *p* = 0.83).

Clinical variables were obtained via chart review. Additional description of the participants and outcomes using different analytic strategies was previously reported (Rose et al., 2015). The experimental protocol was approved by the Stanford University Institutional Review Board #IRB-13899. A parent or legal guardian provided informed written consent and participants were compensated for participation in neurodevelopmental assessments.

### 2.2 Procedures

#### 2.2.1 Diffusion MRI Acquisition, Measures, and Analyses

##### 2.2.1.1 MRI ACQUISITION

MRI data were acquired on a 3T Discovery MR750 scanner equipped with an 8-channel HD head coil (General Electric Healthcare, Little Chalfont, UK). Infants were swaddled and fed and typically remained asleep during the scan. Sedation was not utilized as part of the research protocol and was not utilized for routine pre-discharge MRI. Clinical neuroradiological assessment of the conventional MRI scans were obtained by chart review. High-resolution T1-weighted anatomical images were collected for each subject. A gradient echo 3-plane localizer was used, and an asset calibration was proscribed to utilize parallel imaging. From the 3-plane localizer, T1-weighted images were collected with the following parameters: TE = 8, TR = 3s, FOV = 24 cm, matrix size = 256 × 256, slice thickness 1.0 mm, voxel size = 0.93 x 0.93 x 1mm, NEX = 1. This T1-weighted image was used as a common reference for alignment of the diffusion tensor image (DTI) maps. dMRI data were acquired with a single-shot, spin-echo, echo-planar imaging sequence with a slice thickness of 3.0 mm, a matrix size of 128 × 128, a 90° flip angle, FOV = 20 cm, voxel size = 1.5 mm x 1.5 mm x 3.0mm, TE = 88.8 ms, TR = 8,000, (*b* = 1,000 s/mm^2^). Diffusion was measured along 25 directions, with three b=0 images.

##### 2.2.1.2 DATA PREPROCESSING

Data preprocessing steps, motion correction procedures, diffusion tensor estimation, and individual native space tractography are described below and have been described in our previous work (Dodson et al., 2017; Travis et al., 2017). The T1-weighted images were first aligned to the canonical ac-pc orientation. Diffusion weighted images were pre-processed with open-source software, Vistasoft *(https://github.com/vistalab/vistasoft)* implemented in MATLAB R2012a (Mathworks, Natick, MA). Subjects’ motion during the diffusion-weighted scan was corrected using a rigid body alignment algorithm (Rohde et al., 2004). Each diffusion weighted image was registered to the mean of the three non-diffusion (b=0) images and the mean b=0 image was registered automatically to the participant’s T1-weighted image, using a rigid body transformation (implemented in SPM8, http://www.fil.ion.ucl.ac.uk/spm/; no warping was applied). The combined transform that resulted from motion correction and alignment to the T1 anatomy was applied to the raw data once, and the transformed images were resampled to 2 x 2 x 2 mm isotropic voxels. This step was performed because non-isotropic voxels may bias the tensor fit and distort both tracking and measurements of diffusion properties (Oouchi et al., 2007). Diffusion gradient directions were then adjusted to fit the resampled diffusion data (Leemans and Jones, 2009).

For each voxel in the aligned and resampled volume, tensors were fit to the diffusion measurements using a standard least-squares algorithm, Robust Estimation of Tensors by Outlier Rejection (RESTORE), which is designed to remove outliers at the tensor estimation step (Chang et al., 2005). A continuous tensor field was estimated using trilinear interpolation of the tensor elements. The eigenvalue decomposition of the diffusion tensor was calculated and the resulting three eigenvalues (λ1, λ2, λ3) were used to compute FA, MD (i.e., the mean of λ1, λ2, and λ3), AD (λ1), and RD ((λ2 + λ3)/2) (Basser and Pierpaoli, 1996).

##### 2.2.1.3 HEAD MOTION

All dMRI scans were rigorously assessed for head motion using the following procedures: We quantified the amount of relative head motion (in mm) in each participant by calculating the magnitude of motion correction in voxels (resampled voxel size = 2 mm isotropic) in the x-y-z planes of each volume relative to the prior volume. We then calculated the mean relative motion for each participant. Participants who had mean relative motion less than 0.7mm were retained for analyses (Huber et al., 2019; Yendiki et al., 2014). Using these stringent criteria, 21 of the 66 participants were excluded.

##### 2.2.1.4 TRACTOGRAPHY

Current analyses focused on *a priori selected* white matter tracts that have been previously implicated in processing linguistic information, depicted in **Figure 1**.

Specifically, we identified tracts of a dorsal pathway, the Arc-L (Dodson et al., 2018; Travis et al., 2016) and two bilateral ventral pathways, the ILF-L and ILF-R, and the UF-L and UF-R (Beaulieu et al., 2005; Ben-Shachar et al., 2007; Bruckert et al., 2019a; Dodson et al., 2018; Travis et al., 2016, 2017; Vandermosten et al., 2012). Additionally, we examined the CC-Occ to capture posterior white matter regions associated with language related skills (Dougherty et al., 2007; Dubner et al., 2019; Frye et al., 2008; Hasan et al., 2012; Huber et al., 2019; Odegard et al., 2009) and susceptible to injuries from PT-birth related complications (Back, 2015; Volpe, 2009). Due to previously described limitations of deterministic tractography for segmenting the right arcuate fasciculus, we made the *a priori* decision to exclude the right arcuate fasciculus from our analyses (Catani et al., 2007; Lebel and Beaulieu, 2009; Mishra et al., 2010; Travis et al., 2015a; Yeatman et al., 2011).

**Figure 1.**
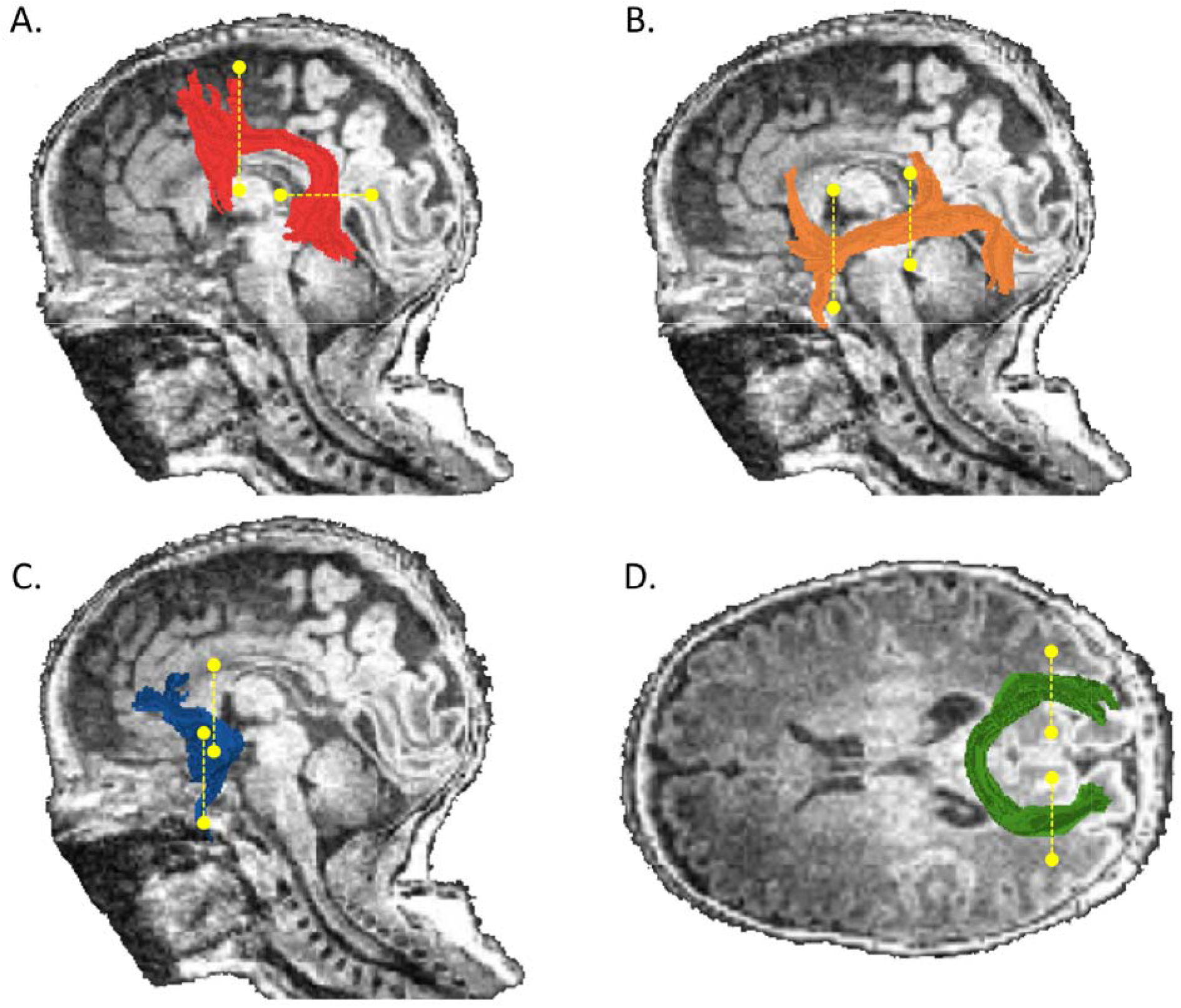
Tractography of selected white matter tracts. Left hemisphere tract renderings are displayed on a mid-sagittal T1 image from a representative participant. Right hemisphere tract renderings not shown. A.) The arcuate fasciculus is shown in red, B.) the inferior longitudinal fasciculus is shown in orange, C.) the uncinate fasciculus is shown in blue. D.) Occipital segment of the corpus callosum rendering is displayed in green on an axial T1 image from the same participant. Dashed lines represent the location of the regions of interest (ROIs) used to segment each pathway from the whole-brain tractogram.

Our dMRI analytic approach was deterministic tractography. In order to maximize sensitivity while taking into account considerable individual variability, particularly at this early stage in brain development, we segmented individual tracts in the native space of each child, then quantified mean diffusivity properties for the tract and also measured diffusivity properties along the trajectory of the tract. We achieved these procedures by performing automatic tract segmentation and quantification using the Automated Fiber Quantification (AFQ; *https://github.com/yeatmanlab/AFQ;* version 0.1) software package and MATLAB 2012a. AFQ uses a three-step procedure to identify each tract in each individual: whole brain tracking, automatic segmentation using anatomically defined template Regions Of Interest (ROIs) in standard (MNI) space warped into native space, and automatic refinement and cleaning (Yeatman et al., 2012b). The AFQ pipeline has been successfully implemented for neonatal images in infants as young as 1 day old (Bruckert et al., 2019b; Langer et al., 2017). To begin the process, the individual’s T1 weighted image is aligned with a template containing the ROIs. We used the default adult template provided in the AFQ software, MNI JHU T2 (Hua et al., 2008; Mori et al., 2005; Wakana et al., 2007). The purpose of this step is to allow placement of the ROIs within the participant’s native space, where fiber tracking is completed. We checked ROI placement in 20% of infants and assured that the ROI placement was accurate. Subsequent AFQ procedures have been described extensively in several previous studies (Dodson et al., 2017; Travis et al., 2015a, 2015b, 2016, 2017; Yeatman et al., 2012b). In the current study, our approach for segmenting dorsal and ventral tracts was similar to Travis *et al*., (Travis et al., 2015a). Procedures for segmenting the posterior corpus callosum were similar to those described in (Dubner et al., 2019). This procedure involves segmenting the corpus callosum into 8 discrete and non-overlapping regions based on the cortical zone where fiber projections terminated (ie., occipital terminations for the posterior corpus callosum), a method that allows for anatomically specific and functionally relevant divisions of the corpus callosum (Dougherty et al., 2007; Huang et al., 2005).

To account for the immaturity of the infant brain, we adjusted the tracking algorithm (Basser et al., 2000; Mori et al., 1999) to perform tracking beginning in voxels from a white matter mask of FA > 0.1 and to stop tracking when voxels dropped below a threshold of FA = 0.08. Individual tractograms representing each fiber group in each subject were visually inspected to confirm that each tract conformed to anatomical norms. We performed this step to ensure that fiber groups did not contain multiple additional streamlines or false positives because of the low threshold needed to perform tracking in the neonatal brain. Tracts that did not conform to anatomical norms were excluded from statistical analyses. After visual inspection, we retained 80% of ILF-L tracts, 73% of ILF-R tracts, 100% of UF-L tracts, 100% of UF-R tracts, 93% of CC-Occ tracts, and 47% of Arc-L tracts across participants. In 14 participants, the Arc-L contained large proportions streamlines that were not consistent with its known anatomy and that were not excluded via automated fiber segmentation and cleaning procedures implemented here. Since the Arc-L conformed to anatomical norms in only (n=16) 53% of participants, we chose to exclude this tract from further analyses.

##### 2.2.1.5 FIBER TRACT QUANTIFICATION: TRACT PROFILES

For each of the tracts, diffusion properties (FA, MD) were calculated at 30 equidistant nodes along a central portion of each fiber tract bounded by the same two ROIs used for tract segmentation. This procedure generated, for every tract and every individual, an FA tract profile that described the variations in FA along the central portion of the tract. At each node, diffusion properties were calculated by taking a weighted average across all streamlines belonging to this tract. Each streamline’s contribution to the average was weighted by the probability that a fiber was a member of the fascicle, computed as the Mahalanobis distance from the tract core (Yeatman et al., 2012b). This procedure minimizes the contribution of fibers located further from the fiber tract core that are more likely to reflect a mixture of gray and white matter or of different tracts, and thereby minimizes the effect of partial voluming on diffusion estimates. This procedure further ensures that we are able to characterize core tract regions that are anatomically consistent across individuals. Tract profiles were then averaged to produce a single mean value for each of the tracts.

##### 2.2.2 Neurodevelopmental assessment

Participants attended the LPCH High-Risk Infant Follow-Up program, during which they were assessed on the Bayley Scales of Infant Development, 3^rd^ Edition (BSID-III) (Bayley, 2006). The target age for assessment was 18-24 months corrected age. One participant who was assessed at 29.5 months chronological age was included because the use of standard scores adjusts for age. We analyzed the composite language standardized score and the expressive and receptive language subtest scaled scores.

#### 2.2.3 Medical complications

To assess the contributions of number of medical complications to outcomes, we generated a composite variable, MedRisk, similar to that used in previous studies (Adams et al., 2018; Marchman et al., 2019, 2016). MedRisk was calculated by summing the presence of common medical complications of PT birth that increase the risk for poor long term cognitive or behavioral outcomes. These medical complications were any degree severity of intraventricular hemorrhage, bronchopulmonary dysplasia, necrotizing enterocolitis, blood culture proven sepsis, patent ductus arteriosus, and retinopathy of prematurity. No participant had seizures or hearing loss, and all but three had hyperbilirubinemia, therefore these common medical complications were excluded from the MedRisk variable. Participants were grouped into low risk (0 risk factors), medium risk (1 or 2 risk factors), and high risk (≥ 3 risk factors).

### 2.3 Statistical analyses

Statistical analyses were conducted using SPSS (version 26.0.0.1, IBM Corp., 2019). Statistical significance was set at *p* < 0.05. The Shapiro–Wilk test was used to assess whether clinical and neurobiological data were normally distributed. With the exception of PMA, data were normally distributed. We chose to do parametric tests for all associations.

Statistical analyses were planned based on the initial sample of 66 participants. With *α* = 0.05 and 1 - β = 0.8, the study was powered to detect an effect size of 0.2 in a multiple regression with 4 predictors. Based on the final sample size of 30 participants with *α* = 0.05 and 1 - β = 0.7, the study was powered to detect an effect size of 0.40 in a multiple regression with 4 predictors.

#### 2.3.1 Demographic and clinical variables

Pearson correlations were computed to evaluate associations between white matter microstructure metrics, GA and PMA.

#### 2.3.2 Mean-tract associations between two-year language outcomes and infant FA and MD

We conducted a series of hierarchical linear regression models to assess associations between composite, receptive, and expressive language scores and mean FA or MD of the a priori selected tracts. PMA and MedRisk scores were entered in the models in the first step to ensure results were related to white matter microstructure metrics rather than participant age at scan. We next assessed the main effects white matter metrics. To explore the pattern of associations across the number of medical complications, we next determined if there was an interaction of MedRisk moderated the prediction of FA to composite language outcome (Hayes, 2017). The variance inflation factor (VIF) was calculated to assess multicollinearity of each model. We considered VIF values less than 10 to indicate that there was no concern for multicollinearity (Field, 2013).

### 2.4 Secondary Analyses

#### 2.4.1 Along-tract profile associations between two-year language outcomes and infant white matter metrics

We calculated partial correlations of the composite language outcome measure and FA or MD values at each location along the tract profile, controlling for PMA. This analysis was conducted in order to confirm that correlations between language and either FA or MD were not obscured due to the use of mean tract measures. The analysis of tract profiles utilized a nonparametric permutation-based method to control for 30 comparisons along the tract (Nichols and Holmes, 2002). This procedure produced a critical node cluster size for each of the candidate tracts (significant cluster size for all tracts was ≥6 locations/nodes). Partial correlations along a tract were considered significant after correction for within tract comparisons if they occurred in a cluster greater than or equal to than the critical cluster size.

#### 2.4.2 Mean-tract associations between language outcomes and infant AD and RD

We conducted secondary analyses to evaluate the associations of AD and RD with composite measures in tracts in which associations were identified between FA or MD and two-year composite language outcomes. This step was achieved by conducting multivariable linear regressions to assess associations between mean AD or mean RD and composite language scores, including PMA and MedRisk in the models.

## 3. RESULTS

### 3.1 Participant Characteristics

Participant characteristics, neonatal medical conditions, and language outcomes are shown in **Table 1**. PMA and GA were weakly negatively correlated (r = −0.186, *p* = 0.33). Six participants had intraventricular hemorrhage on head ultrasound, all of which was grade I. Clinical MRI scans demonstrated no abnormality (n = 16) or minimal abnormality (n = 14), such as subependymal hemorrhage or mineralization.

**Table 1.**
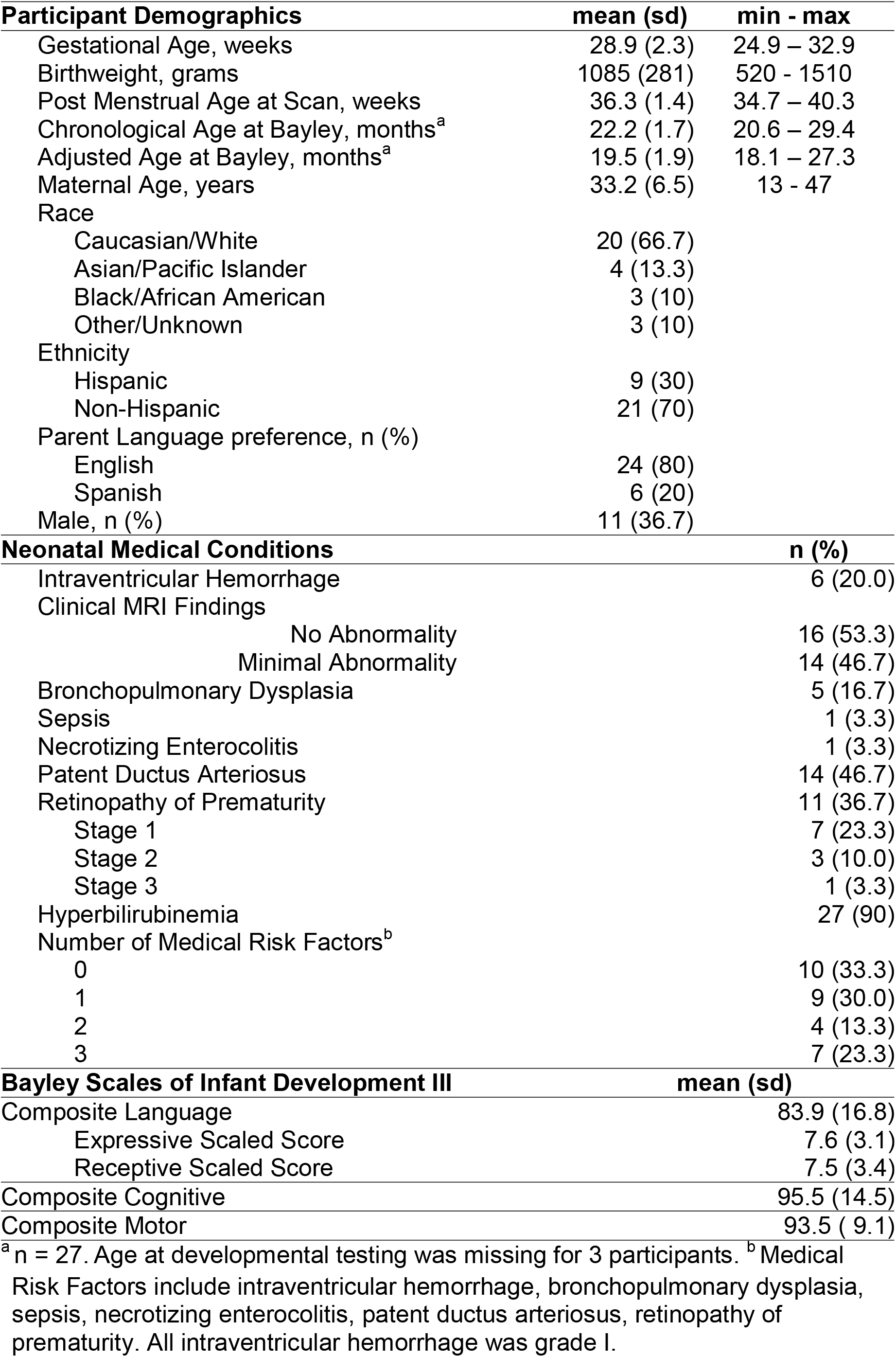
Participant Characteristics (n=30)

| **Participant Demographics** | **mean (sd)** | **min - max** |
| --- | --- | --- |
| Gestational Age, weeks | 28.9 (2.3) | 24.9 – 32.9 |
| Birthweight, grams | 1085 (281) | 520 - 1510 |
| Post Menstrual Age at Scan, weeks | 36.3 (1.4) | 34.7 – 40.3 |
| Chronological Age at Bayley, months<sup>a</sup> | 22.2 (1.7) | 20.6 – 29.4 |
| Adjusted Age at Bayley, months<sup>a</sup> | 19.5 (1.9) | 18.1 – 27.3 |
| Maternal Age, years | 33.2 (6.5) | 13 - 47 |
| Race |  |  |
| Caucasian/White | 20 (66.7) |  |
| Asian/Pacific Islander | 4 (13.3) |  |
| Black/African American | 3 (10) |  |
| Other/Unknown | 3 (10) |  |
| Ethnicity |  |  |
| Hispanic | 9 (30) |  |
| Non-Hispanic | 21 (70) |  |
| Parent Language preference, n (%) |  |  |
| English | 24 (80) |  |
| Spanish | 6 (20) |  |
| Male, n (%) | 11 (36.7) |  |
| **Neonatal Medical Conditions** |  | **n (%)** |
| Intraventricular Hemorrhage |  | 6 (20.0) |
| Clinical MRI Findings |  |  |
| No Abnormality |  | 16 (53.3) |
| Minimal Abnormality |  | 14 (46.7) |
| Bronchopulmonary Dysplasia |  | 5 (16.7) |
| Sepsis |  | 1 (3.3) |
| Necrotizing Enterocolitis |  | 1 (3.3) |
| Patent Ductus Arteriosus |  | 14 (46.7) |
| Retinopathy of Prematurity |  | 11 (36.7) |
| Stage 1 |  | 7 (23.3) |
| Stage 2 |  | 3 (10.0) |
| Stage 3 |  | 1 (3.3) |
| Hyperbilirubinemia |  | 27 (90) |
| Number of Medical Risk Factors<sup>b</sup> |  |  |
| 0 |  | 10 (33.3) |
| 1 |  | 9 (30.0) |
| 2 |  | 4 (13.3) |
| 3 |  | 7 (23.3) |
| **Bayley Scales of Infant Development III** | **mean (sd)** |  |
| Composite Language | 83.9 (16.8) |  |
| Expressive Scaled Score | 7.6 (3.1) |  |
| Receptive Scaled Score | 7.5 (3.4) |  |
| Composite Cognitive | 95.5 (14.5) |  |
| Composite Motor | 93.5 ( 9.1) |  |

<sup>a</sup> n = 27. Age at developmental testing was missing for 3 participants. <sup>b</sup> Medical
Risk Factors include intraventricular hemorrhage, bronchopulmonary dysplasia, sepsis, necrotizing enterocolitis, patent ductus arteriosus, retinopathy of prematurity. All intraventricular hemorrhage was grade I.

### 3.2 Measures of white matter microstructure

Table 2 lists tracts, number of participants in which each tract was identified, and mean (SD) FA and MD. Correlations between tract FA or MD and PMA are shown in Supplementary Table S1. FA of the ILF-L and ILF-R positively correlated with PMA, indicating that as expected FA increased with increasing PMA. MD of the ILF-R, UF-L, and UF-R negatively correlated with PMA, also the expected direction.

**Table 2.** Tract mean FA and MD

| Tract | n | FA mean (SD) | MD mean (SD) |
| --- | --- | --- | --- |
| ILF-L | 24 | 0.19 (0.02) | 1.57 (0.09) |
| ILF-R | 22 | 0.18 (0.02) | 1.51 (0.07) |
| UF-L | 30 | 0.19 (0.02) | 1.40 (0.06) |
| UF-R | 30 | 0.20 (0.02) | 1.39 (0.05) |
| CC-Occ | 28 | 0.33 (0.04) | 1.56 (0.08) |

FA = Fractional Anisotropy, MD = Mean Diffusivity ( $10^{-3}$ mm<sup>2</sup>/sec). ILF = inferior longitudinal fasciculus, UF = uncinate fasciculus, CC-Occ = occipital segment of the corpus callosum. L = Left, R = Right

### 3.3 Hierarchical multivariable linear regression models

**Table 3** shows the results of multivariable linear regression models predicting language outcome at age 2 from FA of the 5 tracts, beyond variation accounted for by PMA. VIF was less than 10 in all regression models. FA of the ILF-L contributed to composite language **(Model 1B)**, explaining an additional 17% of the variance above that accounted for by PMA and MedRisk alone (Model 1A). 42% of the variance in language was accounted for by the model. The interaction term did not contribute additional variance **(Model 1C)**, indicating that the relationship between language outcomes at age 2 and FA of the ILF-L in infancy was similar for children regardless of the number of MedRisk conditions (**Figure 2A**).

**Figure 2.**
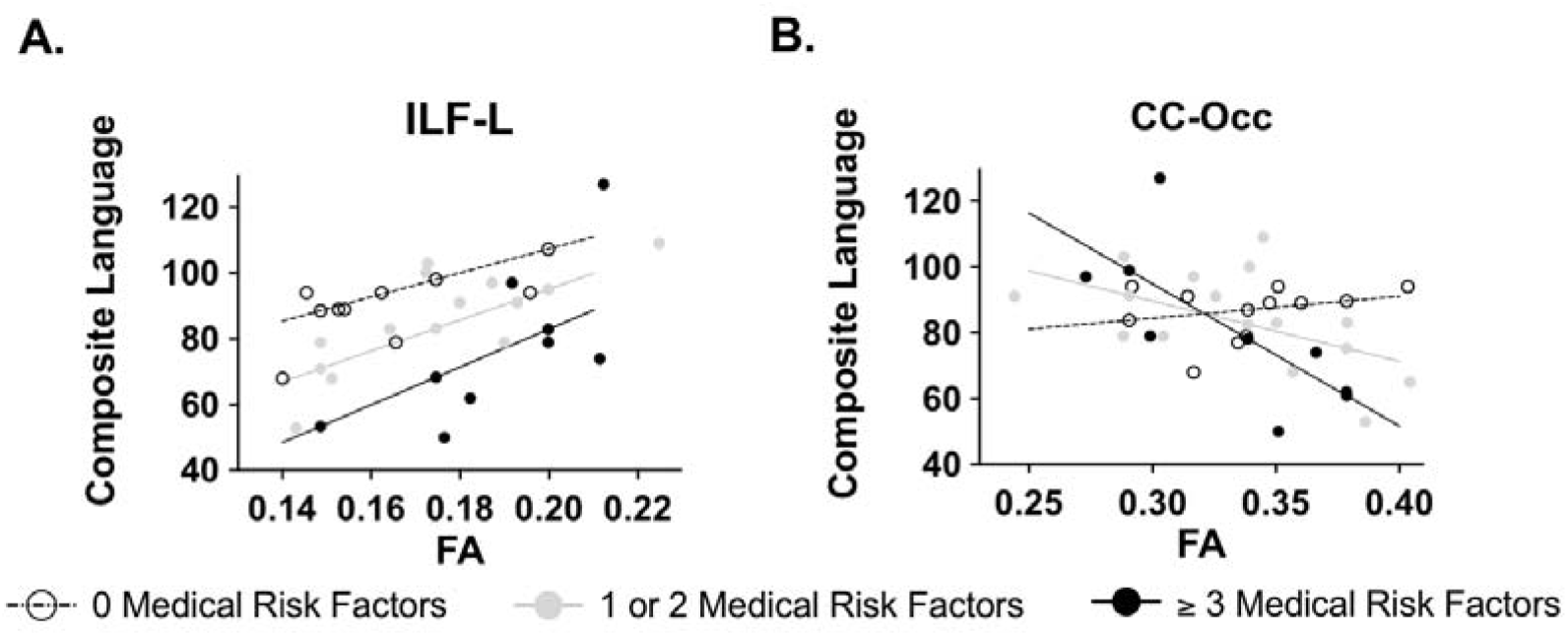
Associations of FA from infant scans obtained prior to NICU discharge and Bayley Scales of Infant Development, 3rd Edition Composite Language Scores, obtained at 18-29 months. Correlations are visualized as scatter plots between mean FA values and composite language values. Predicted model, which is adjusted for post menstrual age at scan, is visualized as regression lines for each MedRisk level. Children with zero medical risk factors are represented with open circles and dashed lines, children with one or two medical risk factors are represented with closed gray circles and solid gray lines, and children with three or more medical risk factors are represented with closed black circles and solid black lines. A) In the left inferior longitudinal fasciculus (ILF-L) the degree of association of ILF-L FA and language was the same regardless of the number of medical complications. B) In the occipital segment of the corpus callosum (CC-Occ), medical complications moderated the association of FA and composite language score. A relation between CC-Occ FA and language at age 2 was found in the participants with 1 or 2 and with 3 or more medical risk factors.

FA of the CC-Occ contributed to composite language (**Model 5B**), explaining an additional 19% of the variance above that accounted for by PMA and MedRisk alone (**Model 5A**). 21% of the variance in language scores was accounted for by the model. The overall model was not significant. The interaction of CC-Occ and MedRisk increased the variance accounted for to 34.1% (**Model 5C**). Moderation analysis indicated that a negative association between language outcomes and FA of the CC-Occ in infancy was found in the participants with 1 or more MedRisk conditions, but not zero MedRisk conditions, as illustrated in **Figure 2B**.

**Table 3.**
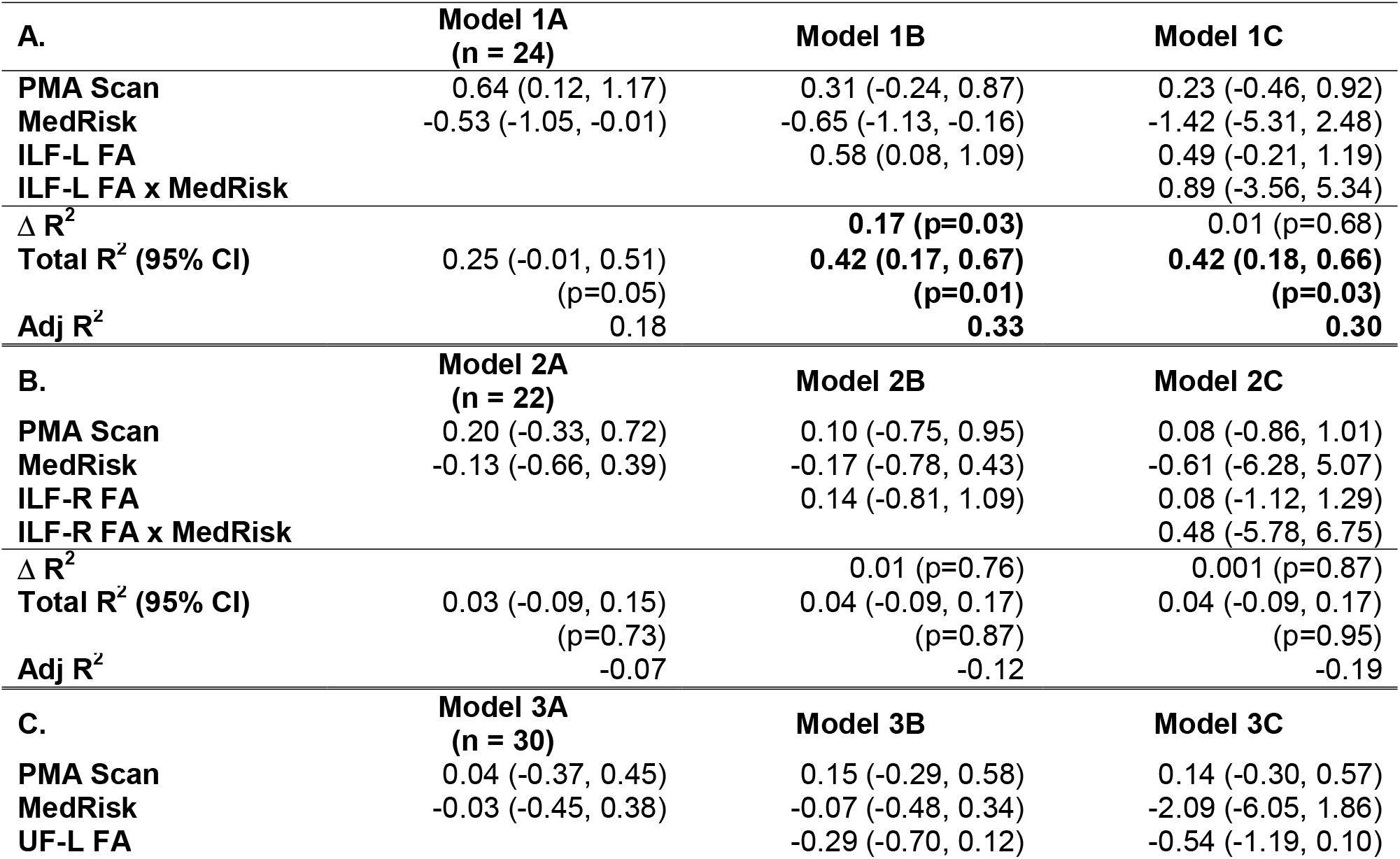
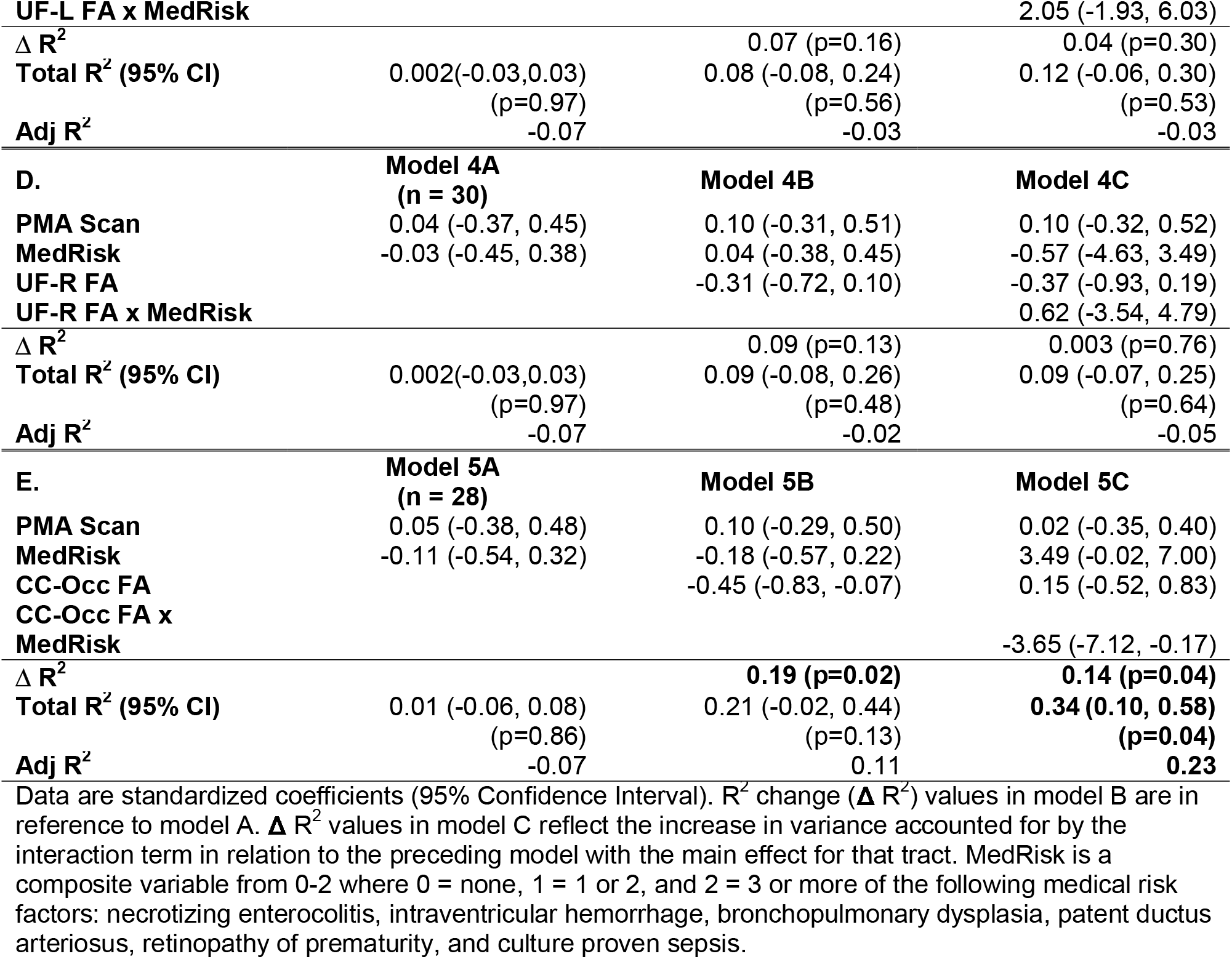
Prediction of language outcome at age 2 years from mean tract-FA of A. left inferior longitudinal fasciculus (ILF-L), B. right inferior longitudinal fasciculus (ILF-R), C. left uncinate fasciculus (UF-L), D. right uncinate fasciculus (UF-R), and E. Corpus Callosum Occipital segment (CC-Occ), beyond covariates of post menstrual age at scan (PMA Scan) and neonatal medical complications (MedRisk).

Supplementary **Table S2** shows the results of multivariable linear regression models predicting language outcome at age 2 from MD of the 5 tracts, beyond variation accounted for by PMA. VIF was less than 10 in all regression models. None of the models were statistically significant.

### 3.4 Secondary Analyses

#### 3.4.1 Along tract profile associations between language outcomes and white matter microstructural metrics

Results of secondary along-tract partial correlations of 2-year language outcomes and along-tract FA and MD, controlling for PMA did not identify additional areas of significant association between FA and composite language scores.

#### 3.4.2 Associations of AD and RD with language

We performed a secondary analysis of multivariable linear regression models predicting language outcome at age 2 from AD and from RD of the 5 tracts, beyond variation accounted for by PMA and MedRisk. None of the models were statistically significant.

## 4. DISCUSSION

In this investigation of 2-year-old children, we evaluated relations between white matter microstructure metrics from infant dMRI obtained prior to hospital discharge and later language outcomes, controlling for PMA at scan and number of neonatal medical complications, and accounting for the moderating effect of number of neonatal medical complications. The major findings of this study were that language scores at age 2 were positively associated with mean FA of the ILF-L and negatively associated with mean FA of the CC-Occ, after accounting for PMA and number of medical complications. We identified a moderating effect of number of medical complications on the association between language and FA in CC-Occ, but not on the ILF-L.

The association between 2-year language outcomes and FA of the ILF-L is consistent with associations in older children born at term (Yeatman et al., 2012a) and preterm (Feldman et al., 2012; Murner-Lavanchy et al., 2018). The ventral pathways, including the ILF, are associated with mapping auditory speech sounds to meaning and simple syntactic structure processing (Hickok and Poeppel, 2007). The associations are consistent with reported deficits in receptive and expressive language skills in children born PT compared with their FT counterparts (Barre et al., 2011).

Language outcomes were negatively associated with FA of the CC-Occ, a white matter region prone to injury in children born preterm (Thompson et al., 2011). The correlation may indicate that FA in posterior periventricular regions indexes general white matter health or injury. In addition, at these very young ages, the posterior corpus callosum may be relevant to language learning, as has been found later in childhood. The observed pattern for negative correlations within the posterior callosal pathway is consistent with reported negative associations between FA and readings skills in older PT children (Dubner et al., 2019) and individuals born FT (Dougherty et al., 2007; Frye et al., 2008; Hasan et al., 2012; Huber et al., 2019; Odegard et al., 2009). Diffusion metrics including FA and MD index multiple tissue properties including but not limited to myelin (Basser and Pierpaoli, 1996; Jones and Cercignani, 2010). Thus, the observed white matter-language relations in the CC-Occ may also reflect variations in axonal properties, such as fiber densities, (Huber et al., 2019) fiber coherence or increased axonal diameter, which have been shown to occur in different proportions across regions of the corpus callosum (Aboitiz et al., 1992). White matter microstructure changes over time as children grow (Lebel and Deoni, 2018). FA may be indexing different axonal properties in infants than in older children. Future studies in larger samples and employing MRI techniques more specific to myelin (Mezer et al., 2013)and analytic approaches for interrogating axonal properties using dMRI data (Rokem et al., 2015; Zhang et al., 2012) are required to clarify the relations between tissue properties across regions of the corpus callosum and later outcomes in children born preterm. The combination of such methods and analytic approaches in future studies may also be important for interrogating the underlying tissue properties contributing the variations in FA and MD at these early stages. Such studies could also help to clarify why 2-year language outcomes were most related to variations in FA but not MD in our sample.

We found that FA of the ILF-L explained additional variance in language beyond number of medical complications. We speculate that the unique contribution of white matter microstructure and the absence of moderation by medical complications in this pathway may relate to the importance of the maturational state of oligodendrocyte precursors relative to the timing and location of oxidative-stress induced brain injuries. For example, prolonged myelination of the ILF (Dubois et al., 2014) may reduce the susceptibility of such regions to injury in the neonatal period. Meanwhile the number of medical complications moderated the tract-language association in the posterior corpus callosum, with greater contribution of FA of the CC-Occ to prediction of language in children with more complications. The posterior corpus callosum myelinates earlier than the ILF (Dubois et al., 2014) and is in a region more prone to injury in the neonatal period. A child with more medical complications in the neonatal period may have greater injury to oligodendrocyte progenitors (Volpe, 2009) in the posterior periventricular region. Our findings suggest that PT children with fewer medical complications may rely primarily on language-related white matter pathways similar to FT children. Children with more medical complications may rely on a different balance of white matter properties in acquiring language, contributing to variation in language development.

The study has several limitations. The sample of neonates whose data contributed to the analyses was modest. Using a stringent threshold for head motion, fewer than half of the scans from the original cohort could be used for analyses. The number of individual tracts included in the analyses was reduced further because of the technical challenges of segmenting the tracts using deterministic tractography. Future studies should enroll a large sample to increase confidence in the results.

We restricted analyses to tracts likely to be associated with language. We did not evaluate the whole brain to determine the specificity of our findings. Future studies should consider assessing the entire brain to confirm the findings described here and to explore whether other areas in the brain are related to language outcomes. We did not have information about other variables associated with language outcomes, such as socioeconomic status (SES), that may contribute to outcomes or may be confounded with other variables. Children with more medical complications may have been born to mothers with lower SES, which may be the true moderator of the relation between FA of the CC-Occ and language. Even for infants with similar brain injury, SES moderates outcomes (Benavente-Fernández et al., 2019). Finally, we used a simplistic measure of neonatal illness. While conditions such as necrotizing enterocolitis, bronchopulmonary dysplasia, sepsis, and intraventricular hemorrhage contribute to hypoxia, ischemia, and inflammation, the exact mechanism by which these conditions lead to changes in white matter metrics is unknown. These major medical complications may directly affect white matter, or they may be a proxy for other disturbances such as prolonged poor nutritional status. Medical complications may also be a marker for other environmental influences associated with altered white matter microstructure, such as increased number of painful procedures (Vinall et al., 2014). These related endogenous or exogenous insults may be the ultimate culprits affecting white matter development.

## 5. CONCLUSION

In summary, a sensitive tractography method identified relations between language skills at age 2 years and infant dMRI white matter metrics from scans obtained prior to hospital discharge. Neonatal medical complications moderated the relations between white matter microstructure metrics in the posterior corpus callosum, suggesting that the white matter properties associated with language outcomes may relate to location and stage of myelination at injury and at the time of scan. Improvements in image acquisition and processing will potentially improve the clinical utility of this approach for identifying early markers of language development. White matter at near-term is one of many possible factors influencing later language, with the post-NICU environment having a large role (Adams et al., 2018). The detectable association between near-term white matter metrics and later language reported here requires further in-depth exploration of the contributors to language development and their interactions in larger samples of preterm infants.

## Data Availability

Anonymized data will be made available upon request to qualified investigators for purposes of replicating procedures and results from the corresponding author.

## Acknowledgments

We thank the children and families who participated in the study; K Yeom for assistance and expertise in reviewing clinical imaging; K Cunanan and K Vitale of the Stanford University Quantitative Sciences Unit for assistance with the statistical approach.

## FUNDING

This work was supported by the National Institutes of Health [NICHD grant RO1-HD069162], and the Health Resources and Services Administration Maternal Child Health Bureau [T77MC09796] to Heidi M. Feldman, PI, the National Institutes of Health [5K99HD084749] and the 2014 Society for Developmental and Behavioral Pediatrics Young Investigator Award to Katherine E. Travis, PI, and the Stanford Maternal Child Health Research Institute Tashia and John Morgridge Endowed Postdoctoral Fellowship to Sarah E. Dubner. This research was also supported by the Chiesi Foundation, Parma Italy; NIH Clinical and Translational Science Award UL1RR025744 for the Stanford Center for Clinical and Translational Education and Research (Spectrum) and for Stanford Center for Clinical Informatics, Stanford Translational Research Integrated Database Environment (STRIDE); Lucile Packard Foundation for Children’s Health; NSF Graduate Research Fellowship grant [DGE-1147470], and by the Mary Baracchi Research Fund, Lucile Packard Children’s Hospital at Stanford.

## Footnotes

1 Abbreviations: AD, axial diffusivity; ANOVA, one-way analysis of variance; AFQ, Automated Fiber Quantification; Arc-L, left arcuate fasciculus; Arc-R, right arcuate fasciculus; BSID-III, Bayley Scales of Infant Development, 3^rd^ Edition; CI, Confidence Interval; CC-Occ, occipital segment of the corpus callosum; dMRI, diffusion magnetic resonance imaging; DTI, Diffusion Tensor Imaging; FA, fractional anisotropy; FT, full term; GA, gestational age; ILF-L, left inferior longitudinal fasciculus; ILF-R, right inferior longitudinal fasciculus; LPCH, Lucile Packard Children’s Hospital; MD, mean diffusivity; MedRisk, number of medical complications; MRI, magnetic resonance imaging; NICU, Neonatal Intensive Care Unit; PMA, post menstrual age; PT, preterm; RD, radial diffusivity; ROI, region of Interest; TEA, term equivalent age; UF-L, left uncinate fasciculus; UF-R, Right uncinate fasciculus; VIF, Variance Inflation Factor

## Notes

### Competing Interest Statement

The authors have declared no competing interest.

### Funding Statement

This work was supported by National Institutes of Health [NICHD grant RO1-HD069162] and Health Resources and Services Administration Maternal Child Health Bureau [T77MC09796] to Heidi M. Feldman, PI, the National Institutes of Health [5K99HD084749] and the 2014 Society for Developmental and Behavioral Pediatrics Young Investigator Award to Katherine E. Travis, PI, and the Stanford Maternal Child Health Research Institute Tashia and John Morgridge Endowed Postdoctoral Fellowship to Sarah E. Dubner. This research was also supported by the Chiesi Foundation, Parma Italy; NIH Clinical and Translational Science Award UL1RR025744 for the Stanford Center for Clinical and Translational Education and Research (Spectrum) and for Stanford Center for Clinical Informatics, Stanford Translational Research Integrated Database Environment (STRIDE); Lucile Packard Foundation for Children's Health; NSF Graduate Research Fellowship grant no. DGE-1147470, and by the Mary Baracchi Research Fund, Lucile Packard Children's Hospital at Stanford.

### Summary of Updates

Methods and results edited. We have elected not to report zero-order correlations between white matter metrics and language outcomes. These correlations are inherently confounded by the developmental age at which infants were scanned. In light of this confound, we revised our analyses to include postmenstrual age at scan (PMA) as a covariate in all analyses. The main results are now reported as independent multivariable linear regressions (Tables 3 and S2).

